# Inactivating mutations in polyadenylation factor CSTF2 disrupt intronic polyadenylation patterns and are associated with intellectual disability

**DOI:** 10.64898/2026.11.10.26367664

**Authors:** Yaroslav Kainov, Patricia A. Foley, Rebecca Igbokwe, Mary-Alice Abbott, Ingrid M. Wentzensen, Andres Ramos, Tim Hubbard, Cristina Dias

## Abstract

Alternative polyadenylation contributes extensively to transcript diversification and is particularly prominent during neuronal differentiation. *CSTF2* encodes an RNA-binding subunit of the cleavage stimulation factor complex and has previously been associated with X-linked intellectual disability through a single Asp50Ala missense variant with increased RNA-binding activity. Here, we identify seven rare *CSTF2* variants in ten hemizygous male probands with intellectual disability, global developmental delay and/or autism spectrum disorder, including missense, nonsense, splice-site, and frameshift variants.

We functionally assessed selected variants by ectopic expression in CSTF2-depleted HEK293T cells, followed by total and 3′-end-proximal RNA sequencing. Phe19Leu, Gln543Ter and Leu555Ser markedly impaired CSTF2 function. These variants predominantly reduced the usage of strong, evolutionarily conserved intronic polyadenylation sites with prominent GU-rich sequences, while increasing the usage of downstream sites within terminal 3′UTRs. In contrast, the previously reported Asp50Ala variant produced an opposing pattern, with preferential activation of intronic polyadenylation sites, consistent with its increased RNA-binding activity. These findings data provide genetic and functional support for *CSTF2* as an X-linked intellectual disability, identify inactivating CSTF2 variants as a disease mechanism, and imply that both reduced and increased CSTF2 activity can result in neurodevelopmental disorder.

## Introduction

In eukaryotes, most protein-coding transcripts undergo a series of mRNA processing steps, including 5′ capping, splicing, and 3′ cleavage and polyadenylation (“polyadenylation” hereafter)^1^. These processes are fundamental for efficient gene expression, with polyadenylation playing a key role in transcription termination, mRNA export, stability and translation^2^. Human genes typically contain multiple polyadenylation sites located within 3′ untranslated regions (UTRs) or introns, resulting in transcript diversification through alternative polyadenylation^3^. Pronounced shifts in alternative polyadenylation patterns are observed during stem cell differentiation and tissue development as well as in cancer cells after malignant transformation^4,5^. Cell differentiation and decrease in proliferation rates is known to be associated with the usage of the distal polyadenylation sites leading to 3’UTR lengthening^6,7^, with differentiated neurons exhibiting exceptionally long 3′UTRs^8^. These findings implicate alternative polyadenylation as an important layer of post-transcriptional control in cell and tissue specification^9,10^.

Polyadenylation is mediated by distinct polyadenylation signals recognized by multi-protein complexes^11,12^. The canonical AATAAA hexamer (PAS-hexamer) upstream of the cleavage site is recognized by the cleavage and polyadenylation specificity factor (CPSF), whereas the cleavage stimulation factor (CSTF) complex binds GU-rich sequences downstream^13^. Additional RNA-binding proteins and auxiliary elements modulate this process, influencing polyadenylation site selection and polyadenylation efficiency^14^. The CSTF complex comprises CSTF1 (CSTF50), CSTF2 (CSTF64), and CSTF3 (CSTF77). As an RNA-binding protein CSTF2 interacts directly with GU-rich elements through its N-terminal RNA recognition motif (RRM) domain^15^ and has established roles in immune cell maturation, pluripotency, and embryonic development^16–18^. We have previously identified a composite RNA-binding site in the RRM yeast orthologue Rna15, which is conserved in mammals and supports a general role for CSTF2 in recognising GU- and U-rich sequences during pre-mRNA 3′-end processing^19^. Expression of numerous nervous-system-specific CSTF2 isoforms further points to a role in neuronal differentiation and function^20^.

The *CSTF2* gene is highly constrained against loss-of-function variation in humans (pLI=1 and LOEUF = 0.318) and was predicted to be associated with monogenic human disorder in a systematic analysis of human chromosome X genes^21^. Previously, P. Grozdanov *et al.* identified and characterized the Asp50Ala variant (chrX:100077251A>C, GRCh38.p12) in the RRM domain of CSTF2, which co-segregated with non-syndromic intellectual disability in an affected family^22^. To date, this remains the only described pathogenic variant in *CSTF2*, limiting its diagnostic utility; for example, *CSTF2* is currently classified as an “amber” gene in PanelApp^23^ (indicating limited but suggestive evidence for disease association) and a gene with limited association with neurodevelopment disorders in SysNDD^24^.

Here, we identified and characterised novel likely pathogenic variants in *CSTF2* in probands with intellectual disability and additional features. Using molecular modelling and overexpression assays, we show that these variants likely cause a loss-of-function phenotype, distinct from that of the previously reported Asp50Ala variant, significantly broadening the molecular and clinical spectrum of CSTF2-related disorders.

### Rare Phe1GLeu variant in site II of CSTF2 RNA binding domain observed in proband with global developmental delay

In the GEL 100kGP Rare Diseases dataset^25^, we identified an undiagnosed male proband with a previously uncharacterised Phe19Leu de novo variant in CSTF2 RRM domain (chrX:100820471:T:C (GRCh38)). In contrast with previously described normal motor milestones and mild intellectual disability in a family with an Asp50Ala variant^22^, this proband presented with global developmental delay, intellectual disability and short stature (Supplementary table 1). The Phe19Leu variant was not observed in gnomAD v4.1.0, has a high CADD v1.7 PHRED score of 27.9, and maximum AlphaMissense score of 1.0. Notably, while the Asp50Ala variant was reported to result in a small (∼2-fold) increase in RNA binding^22^, we have previously shown that mutation of Tyr21, the residue corresponding to Phe19 in the yeast orthologue RNA15, to Alanine leads to a significant 10-fold decrease in RNA-binding affinity^19^. The important role of the aromatic sidechain in the Phe19 position, which is conserved from Arabidopsis to human (Fig 1A), in RNA binding is supported by structural and biophysical data in the yeast and human protein^19,26,27^. In yeast, the high-resolution structure of RNA15 in complex with the cognate RNA shows that Tyr21 stacks on an RNA base. This stacking is maintained in the larger yeast multi-protein RNA recognition complex, as shown by NMR^28^. In humans, this residue is conserved, and our Alphafold3^29^ prediction for the human CSTF2 RRM protein in complex with a GU-rich RNA also shows prominent stacking interaction of Phe19 with an RNA base (Fig 1B). Upon *in silico* mutation of Phe19 to Leu, this interaction is lost (Fig 1B). We therefore hypothesised that the Phe19Leu variant has a “loss-of-function” molecular phenotype which potentially contributes to a more severe neurodevelopmental phenotype.

**Figure 1:**
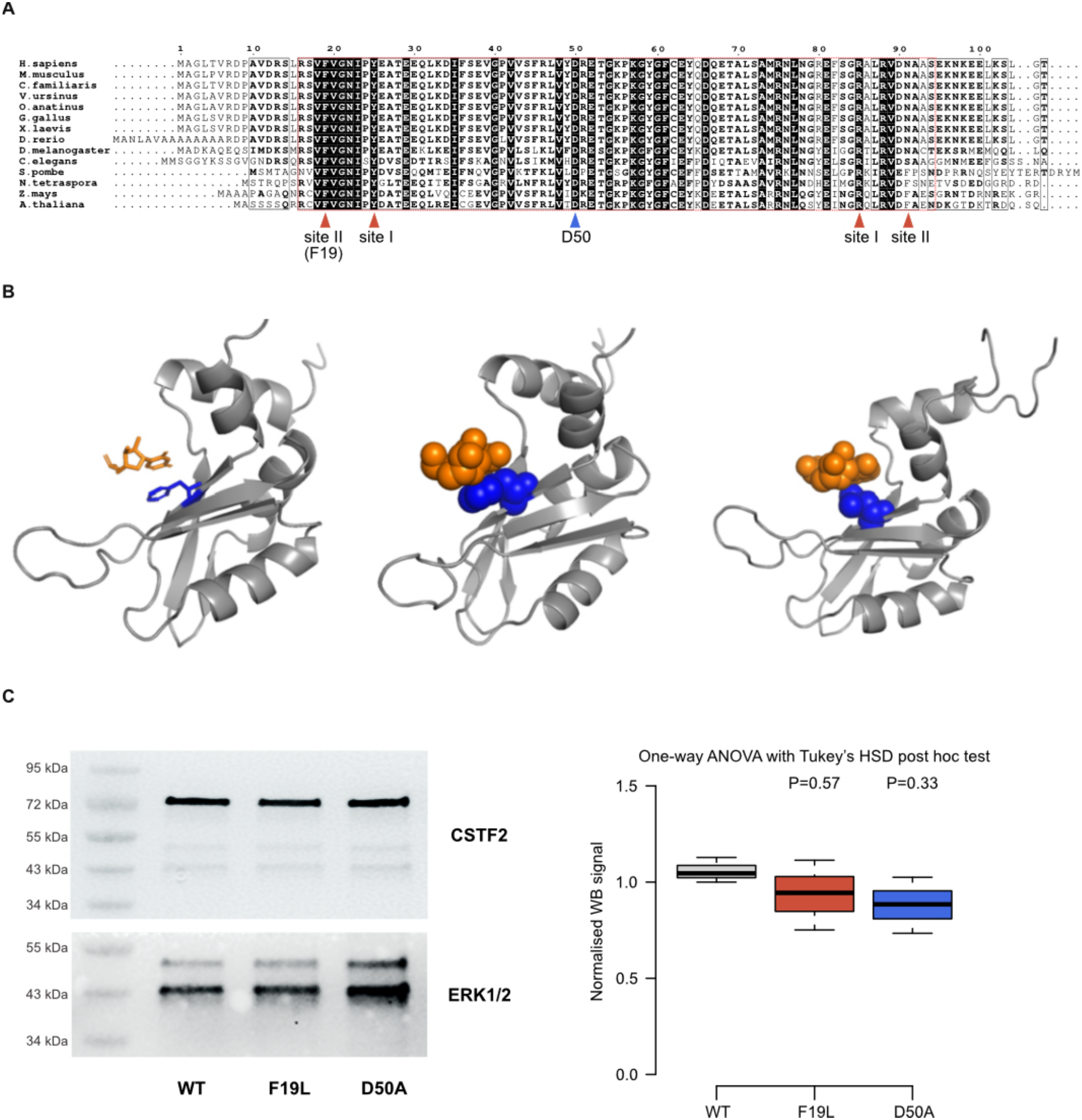
The Phe1GLeu variant occurs in the functionally important site II of the CSTF2 RRM domain and is predicted to abrogate RNA binding. **A -** CLUSTAL alignment of *CSTF2* orthologs showing amino acid conservation in the RRM domain. Note strong evolutionary conservation of the Phe19 residue in site II and at Asp50, where a pathogenic Asp50Ala variant has previously been identified **B -** *(Left and centre)* The aromatic ring of phenylalanine 19 in the human CSTF2 protein stacks against the closest base in the cognate RNA in an Alphafold3 model of the complex. The protein backbone is shown as a silver cartoon, while the RNA nucleotides and the protein tyrosine residue are highlighted in orange and blue, respectively, and rendered as sticks or spheres, to better highlight direct contacts *(Right).* This interaction is largely lost when Phe19 is mutated to leucine - the same colour coding and representation are used. The models are displayed using Pymol ^53^. **C -** Left: immunoblot analysis of CSTF2 variants effect on protein stability. ERK1/2 is included as a loading control. Right: CSTF2 protein expression was quantified from three independent experiments (the box bounds represent the first and the third quartiles and the thick black lines at the bottom of the boxes show the medians).

### Overexpression of Phe1GLeu CSTF2 dysregulates polyadenylation

To experimentally characterise the impact of the CSTF2 Phe19Leu mutation on polyadenylation in a cellular context, we transiently overexpressed human wild-type (WT) CSTF2 and Phe19Leu and Asp50Ala mutants in HEK293T A2 cells with knock-down of endogenous CSTF2 and performed whole transcriptome analysis by RNA-seq. We did not observe significant changes in the stability of CSTF2 mutants after overexpressing their V5-tagged ORFs (Fig 1C). Using IPAFinder^30^ to quantify and compare the relative usage of intronic polyadenylation sites in total RNA-seq data (see Methods), we identified pronounced changes in usage of intronic sites in the case of the Phe19Leu mutant overexpression in comparison to wild-type CSTF2, but not in overexpression of Asp50Ala (Fig 2). Overall, 194 intronic polyadenylation sites were significantly suppressed in Phe19Leu samples in comparison with WT, and only 36 intronic sites were activated (Fig 2A) in keeping with the predicted “loss-of-function” nature of the variant. Interestingly, a significantly different pattern of changes was observed in case of the Asp50Ala mutant, with 42 intronic sites being significantly activated and 4 repressed in comparison with WT (Fig 2A), supporting previous observations of the Asp50Ala variant having a molecular “gain-of-function” effect^22^. Principal component analysis (PCA) and hierarchical clustering of relative usage of significantly regulated intronic polyadenylation events distinctively separate Asp50Ala and Phe19Leu samples from WT suggesting opposite functional effect of these mutations on CSTF2 polyadenylation activity, in agreement with an opposite effect in RNA binding (Fig 2B and 2C). Of note, similar but less pronounced effect on polyadenylation sites in 3’UTRs was detected using DaPARS tool^5^ (see PCA and clustering in supp. Fig S2,S3). Moreover, of 19 intronic polyadenylation sites that were dysregulated by both mutants (Fig. 2D), most (15 sites, 79%) were suppressed by Phe19Leu and activated by Asp50Ala (Fig. 2E).

**Figure 2:**
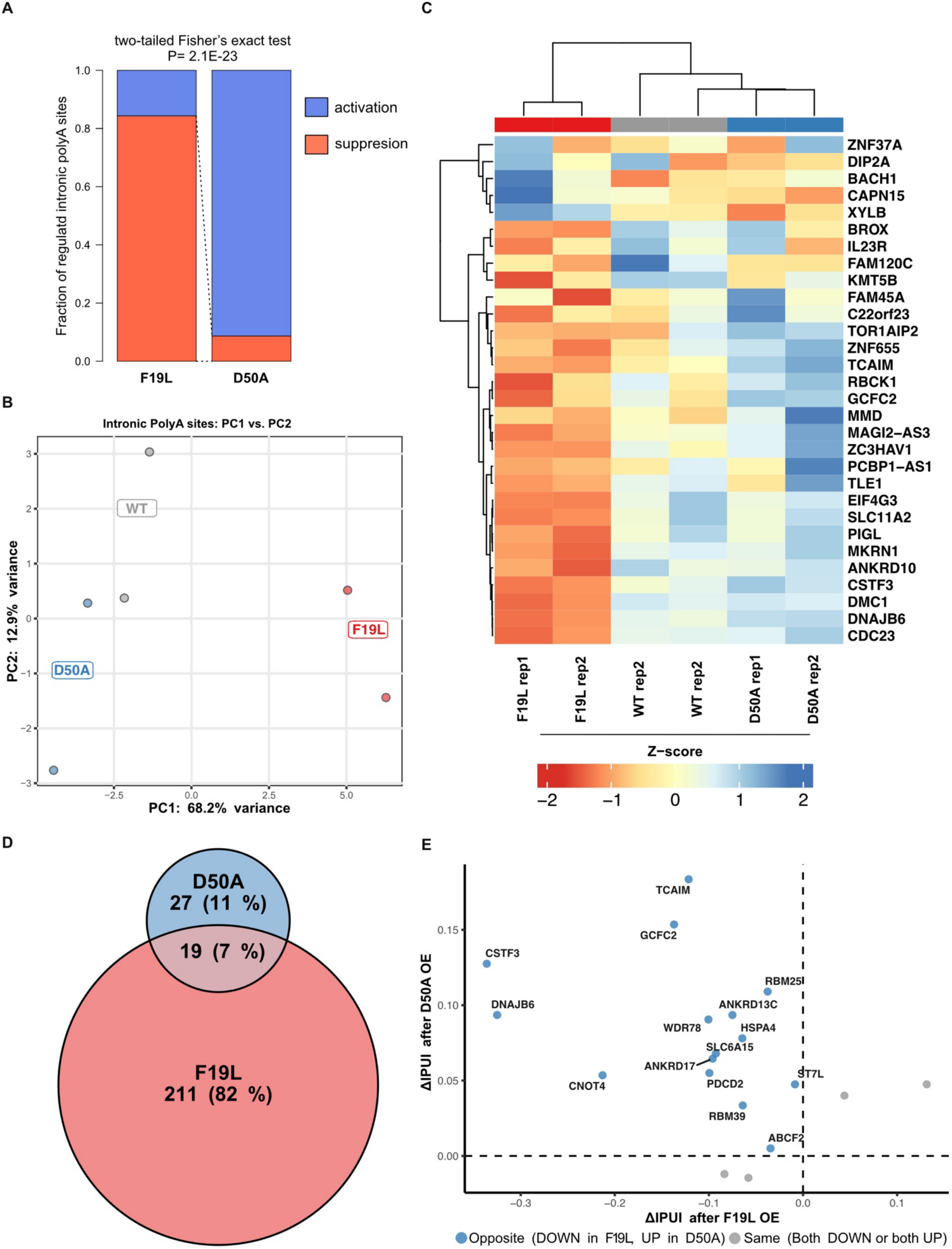
Phe1GLeu CSTF2 variant suppresses intronic polyadenylation in contrast to Asp50Ala. **A** - Overexpression of the Phe19Leu (F19L) CSTF2 ORF leads to strong suppression of intronic polyadenylation compared with the with the Asp50Ala (D50A) variant (two-tailed Fisher’s exact test comparison of all significant intronic polyadenylation events in comparison with WT CSTF2 IPAFinder BH-adjusted p < 0.05). **B** - Principal component analysis (PCA) of the 100 most variable intronic polyadenylation events significantly dysregulated by overexpression of either Asp50Ala CSTF2 (D1, D2) or Phe19Leu CSTF2 (F1, F2) ORFs compared with the with WT CTSF2 ORF in HEK 293T A2 cells (IPAFinder BH-adjusted p <0.05). Samples were projected onto the first two principal components, with group centroids as labelled rectangles calculated as the mean PC coordinates per condition. **C** - Heatmap of the top 30 most variable intronic polyadenylation events significantly dysregulated (IPAFinder BH-adjusted p <0.05) by overexpression of either Asp50Ala CSTF2 or Phe19Leu CSTF2 in comparison with WT CTSF2. Rows and columns were hierarchically clustered using correlation distance and Ward’s D2 linkage (pheatmap, R). PDUI values were scaled by row Z-score and visualized with a red–white–blue colour scheme. Note the opposite effects of Phe19Leu and Asp50Ala variants on intronic polyadenylation. **D** - The proportional overlap (Euler diagram) of significantly dysregulated intronic sites after F19L and D50A CSTF2 **E** – The four-quadrant scatter plot showing directional concordance of shared significantly dysregulated intronic sites from D (19 genes). Discordantly regulated genes highlighted blue. Note striking discordance (p=1.2e-06, one-tailed exact binomial test with 25% fraction assumption) in regulation between F19L and D50A CSTF2 variants.

A striking example of such regulation is the conserved intronic polyadenylation site in intron 3 of the *CSTF3* (Cleavage Stimulation Factor Subunit 3) gene, a well-known target of CSTF2 and a part of an autoregulatory feedback loop^31^. Overexpression of Phe19Leu CSTF2 leads to a decrease of intronic polyadenylation of the *CSTF3* intronic site (Fig 3A), manifested as a marked decrease of intronic reads (almost undetected by IPAFinder in both Phe19Leu replicates) and pronounced increase in expression of the full-length transcript (Fig 3B). Conversely, overexpression of the Asp50Ala mutant leads to increased expression of short intronically polyadenylated isoform and decreased expression of full-length functional *CSTF3* isoform (Fig 3A-B). Similarly, in the *ST7* gene (Suppression Of Tumorigenicity 7), CSTF2 mutants opposingly affect polyadenylation at two alternative 3’end terminal exons, with the proximal one being less often used when putative “loss-of-function” Phe19Leu mutant is overexpressed (Fig 3C).

**Figure 3:**
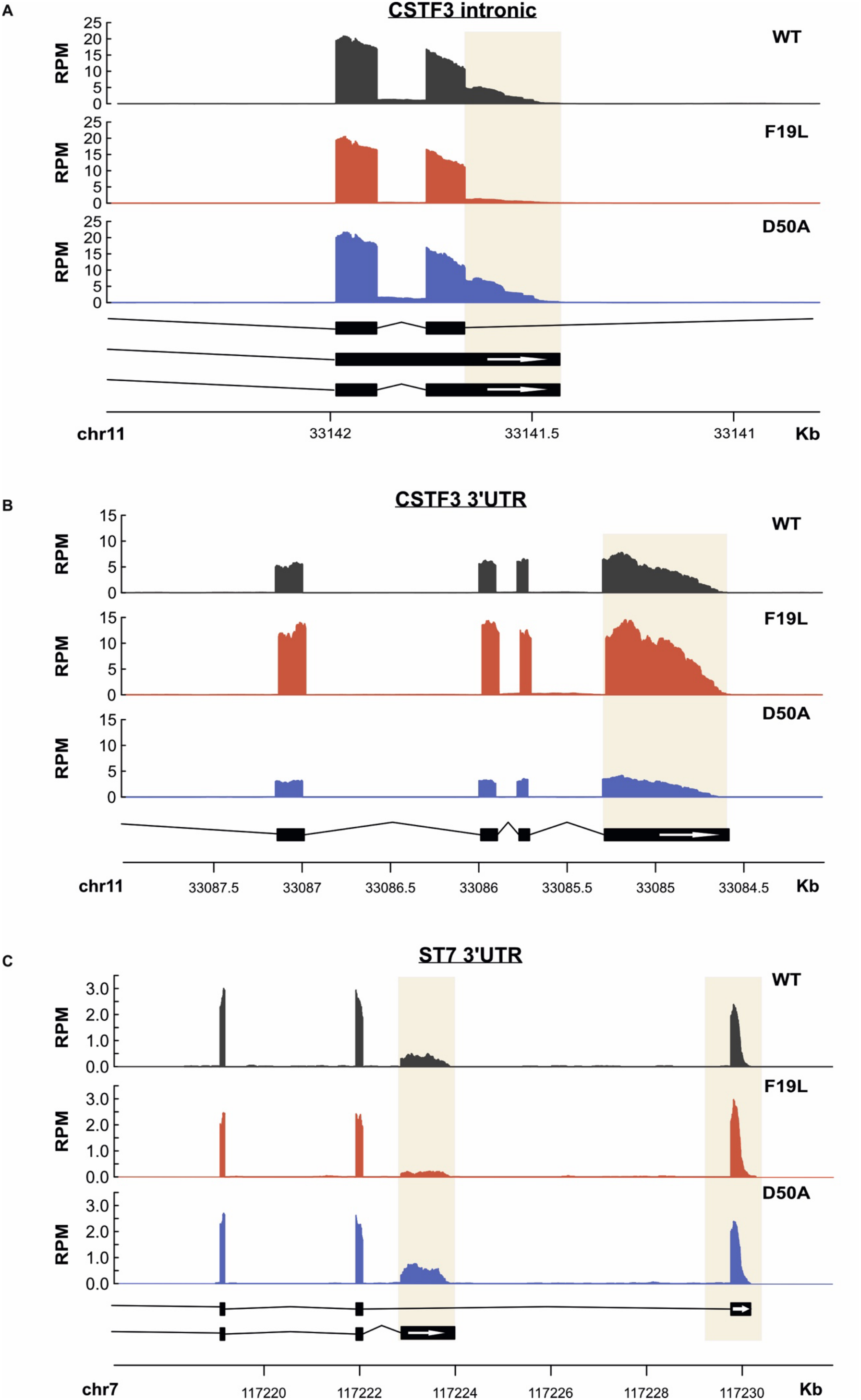
Examples of dysregulated intronic polyadenylation events following overexpression of CSTF2 variants containing ORFs. **A -** decrease of intronic polyadenylation of the *CSTF3* intronic site with overexpression of the F19L mutation (red track) relative to the wild-type (black track), and increase of polyadenylation with overexpression of the D50A mutant (blue track). **B -** pronounced increase in mRNA expression of the 3’ UTR with over expression of F19L, vs. decrease with overexpression of D50A. **C –** Decrease of intronic polyadenylation and increase of 3’ UTR polyadenylation of the *ST7* gene with overexpression of the F19L mutant, and opposing effect of the D50A mutant. Read-per-million (RPM)-normalized RNA-seq coverage plots are shown; white arrows indicate terminal exons additionally highlighted by beige rectangles.

### Inactivating variants in CSTF2 supress intronic polyadenylation

In order to identify other potentially pathogenic inactivating variants in *CSTF2* we looked for rare nonsense, frame-shift, splice site and predicted-to-be-damaging missense variants in functional domains of CSTF2 in hemizygous male probands presenting with intellectual disability, global development delay and/or autism spectrum disorder in GEL 100kGP Rare Diseases, NHS genomic medicine service (GMS), SPARK^32^ cohorts and through GeneMatcher^33^. We identified an additional six novel variants in nine probands, including three missense, one nonsense, one splice site and one frameshift variants (Fig 4A, Supplementary Table 1). Across all individuals with *CSTF2* variants, in addition to global developmental delay and/or intellectual disability (7/10), individuals presented with delayed speech and language development (9/9), autistic behaviours (7/9), feeding difficulties (4/7), motor coordination difficulties (3/7), behavioural abnormalities (3/6), and ADHD (2/5). Developmental regression was reported in two individuals, other features included sleep disturbances, ocular/vision abnormalities, hypotonia, and endocrine abnormalities. We selected all missense variants and stop gain variant Gln543ter, located in the last coding exon and potentially producing a truncated protein, for further functional studies.

**Figure 4:**
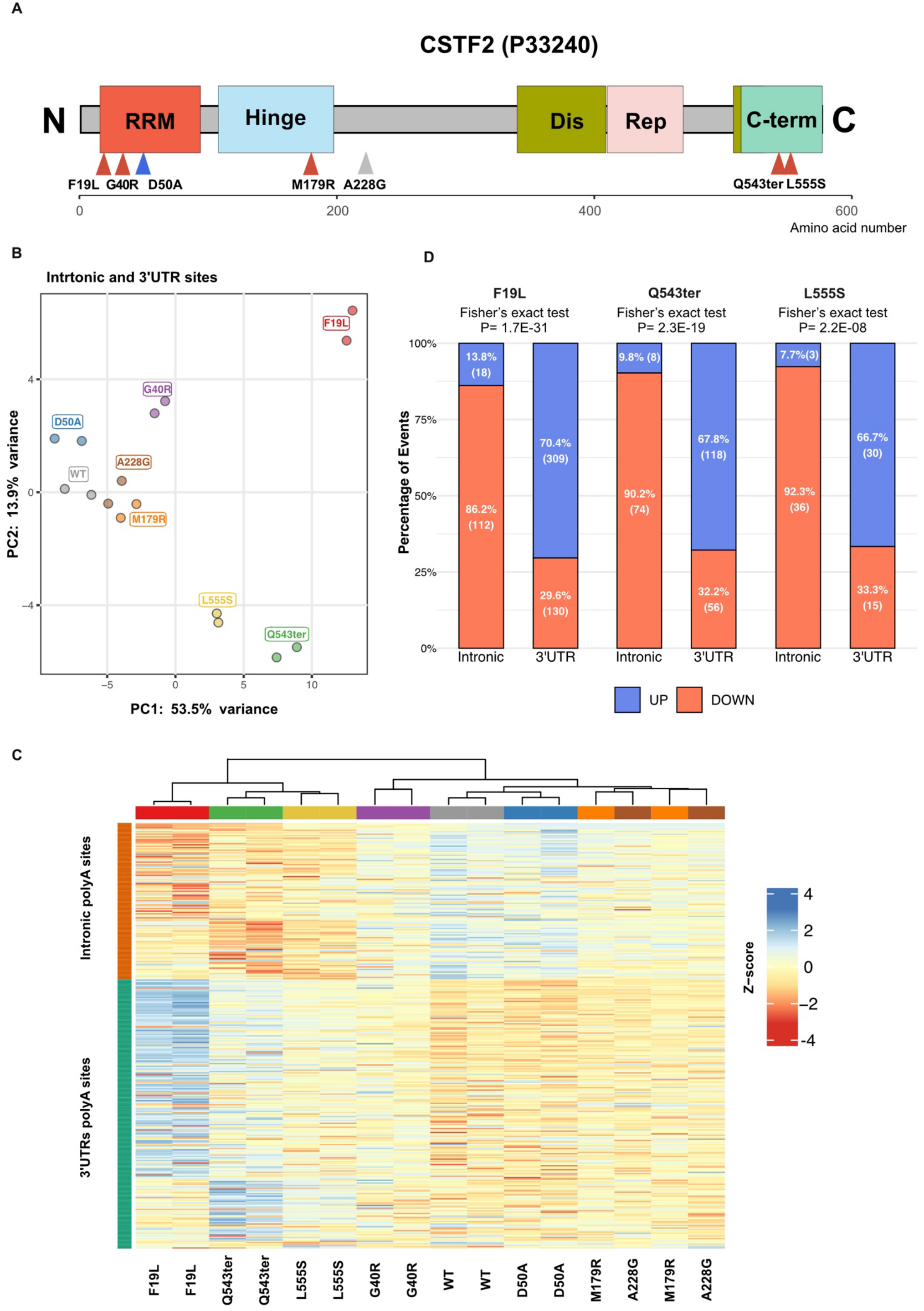
Inactivating CSTF2 variants overexpression supresses intronic and activates 3’UTR polyadenylation sites. **A** - Annotated domain structure of human CSTF2 (UniProt P33240, 577 amino acids). Indicated are the RRM (PFAM: RRM_1 - PF00076), the Hinge (“Hinge”, PFAM: CSTF2_hinge - PF14327), predicted disordered region (“Dis”), the 12× MEARA repeats (“Rep”), and the C-terminal domain (“C-term”, PFAM: CSTF_C - PF14304). Triangles indicate positions of the variants reported here (red), previously reported variant (blue), and population control (grey). **B** – Principal component analysis (PCA) of activity of all polyadenylation sites significantly dysregulated in at least one of seven comparisons with wild-type (WT) CSTF2 in HEK293T A2 cells (Benjamini–Hochberg-adjusted P < 0.05 and |log₂FC| ≥ 1). Samples were projected onto the first two principal components, with group centroids calculated as the mean PC coordinates per condition. Percentages on the axes indicate the variance explained by each principal component. **C** – Heatmap of the same differentially regulated polyadenylation sites. Normalised log₂ CPM values were scaled by row Z-score and visualized with a red–white–blue colour scheme. Sites were grouped by genomic annotation (intronic, 3′UTR and other) without row clustering, while samples were hierarchically clustered using correlation distance and Ward’s D2 linkage (pheatmap, R). Note the opposite effects on intronic and 3’UTR polyadenylation after Phe19Leu (F19L), Gln543ter (Q543ter) and Leu555Ser (L555S) CSTF2 ORFs overexpression. **D** - Overexpression of the Phe19Leu (F19L), Gln543ter (Q543ter) and Leu555Ser (L555S) CSTF2 ORFs leads to opposite effect on intronic and polyadenylation sites (two-tailed Fisher’s exact test comparison intronic site versus 3’UTR sites).

To functionally characterise newly identified *CSTF2* variants we overexpressed them in HEK293T A2 cells as previously, alongside Ala228Gly, a variant predicted-to-be-benign and observed in the general population (gnomAD v4.1.0 AF = 0.003763, AlphaMissense score = 0.068, CADDv1.7 PHRED =13.3), and performed 3’end proximal RNA-sequencing allowing to more precisely analyse polyadenylation shifts. In line with our previous data, we detected a distinct strong global shift in polyadenylation in the Phe19Leu variant (Fig 4B,C). Another RRM domain variant Gly40Arg (absent from gnomAD, AlphaMissense score = 1.0, CADDv1.7 PHRED =35) exhibited a moderate shift in polyadenylation in similar direction. The Met179Arg variant (absent from gnomAD, AlphaMissense score = 1.0, CADDv1.7 PHRED =25.5) located in a Hinge domain (Pfam: PF14327, *CSTF2_hinge*) did not produce a notable shift in polyadenylation activity in our assay. Interestingly two variants located in the conserved C-terminal domain (Pfam:PF14304, *CSTF_C*) responsible for critical interactions with the polyadenylation machinery^34^- Gln543ter and Leu555Ser - also strongly affected polyadenylation (Fig 4B,C). Both variants are predicted to disrupt interaction function of the C-terminal domain: Gln543ter inducing truncation that is predicted to produce an unfolded domain, and Leu555Ser by substituting a polar amino acid to a core non-polar position of the evolutionary conserved motif.

### CSTF2 variants affect intronic polyadenylation at GU-rich evolutionary conserved polyadenylation sites

To further understand the variants’ functional effect on polyadenylation and potential molecular mechanisms we focused on directionality of dysregulation and the features of polyadenylation sites affected by Phe19Leu, Gln543ter and Leu555Ser variants. All these mutants predominantly suppressed intronic polyadenylation sites, but activated polyadenylation in 3’UTR regions at the ends of genes (Fig 4D). At the individual gene level, we identified that suppression of intronic sites often directly co-occurred with higher combined signal from the 3’UTR polyadenylation site in the same gene (Fig 5A, upper left quadrant of plots) suggesting that inactivating CSTF2 mutations induces a shift from short intronic to full length isoforms.

**Figure 5:**
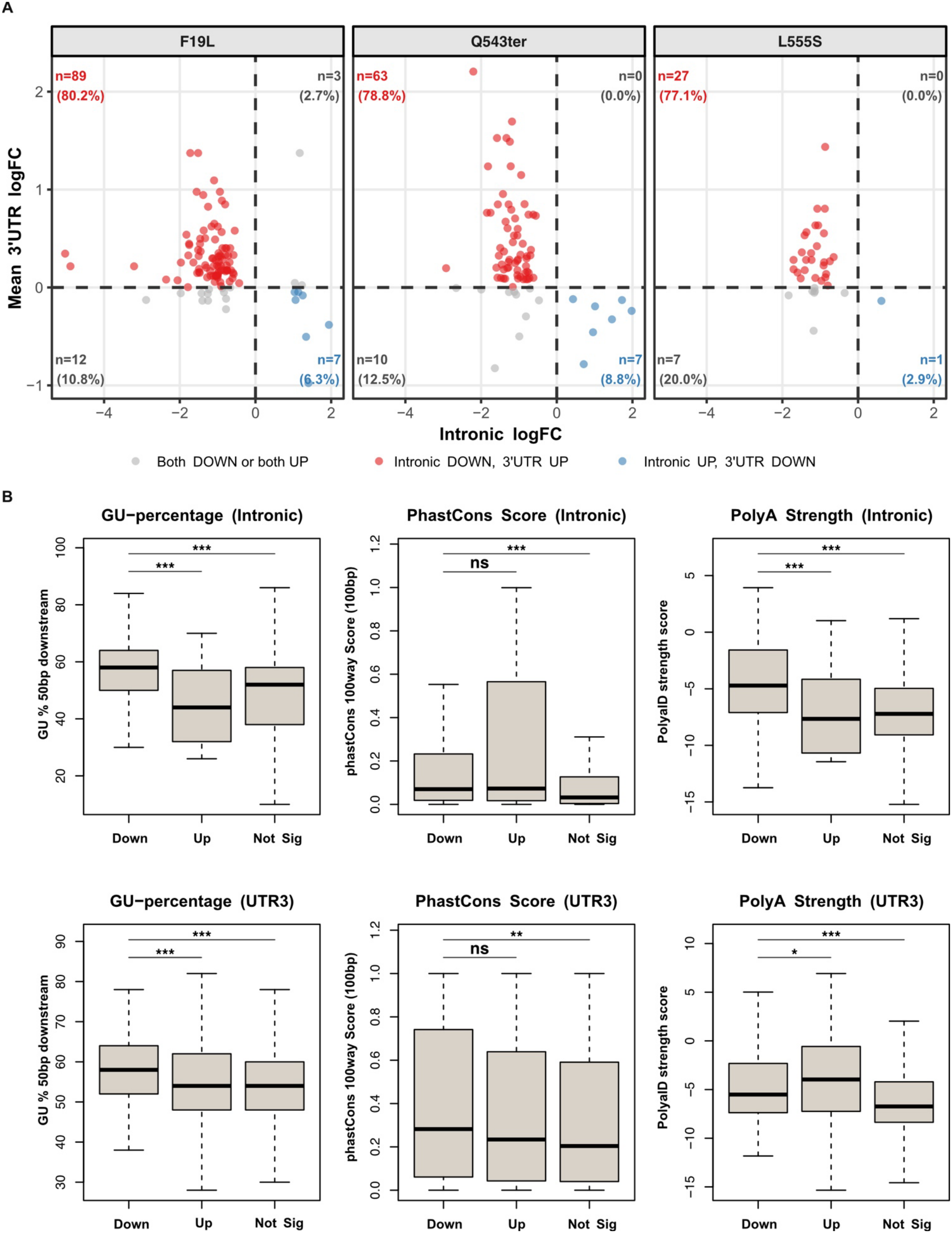
Inactivating CSTF2 variants suppress polyadenylation at strong evolutionary conserved GU-rich polyadenylation sites. **A** – The four-quadrant scatter plot showing upregulation of polyadenylation in 3’UTRs of the same gene where intronic polyadenylation is suppressed for Phe19Leu (F19L), Gln543ter (Q543ter) and Leu555Ser (Leu555Ser) variants. **B** - Boxplots comparing PhastCons conservation scores, poly(A) site strength, and GU percentage 50 bp downstream of the site across down-regulated, up-regulated, and non-significant polyadenylation sites (for the union of sites for for Phe19Leu (F19L), Gln543ter (Q543ter) and Leu555Ser (L555S) variants (BH-adjusted P<0.05))stratified by genomic region (intronic and 3’UTR), with asterisks indicating pairwise Wilcoxon rank-sum test significance relative to the down-regulated sites (* P<0.05, ** P<0.01, *** P<0.001, ns = not significant).

CSTF2 is known to bind to GU-rich elements downstream of polyadenylation sites. Our analysis showed that both intronic and 3’UTR sites that are suppressed by Phe19Leu, Gln543ter and Leu555Ser variants have significantly higher fraction of GU-nucleotides than activated and non-regulated sites (Fig 5B) suggesting its direct effect on this group of sites through impaired binding. Additionally, intronic suppressed sites are predicted to be stronger than all other sites and more evolutionary conserved that non-regulated ones (Fig 5C), with PolyA strength defined as a predicted quantitative log-odds score to measure relative polyadenylation site usage and cleavage strength^35^. A similar but less pronounced trend was observed for 3’UTR suppressed sites in comparison with non-regulated ones (Fig 5B).

Thus, inactivating CSTF2 variants dysregulate polyadenylation by predominantly suppressing a strong evolutionary conserved set of intronic sites.

## Discussion

Here we identified loss-of-function variants in polyadenylation factor CSTF2 in hemizygous probands with intellectual disability, global developmental delay and autism spectrum disorder. For a subset of missense variants, we showed that they dysregulate polyadenylation by impairing functions of evolutionary conserved functional domains of CSTF2. This functional evidence implicating CSTF2 in RNA cleavage and polyadenylation provides a plausible molecular mechanism for the neurodevelopmental phenotypes observed in the cohort, particularly speech and language delay, intellectual disability and autistic features. Notably, the previously reported familial Asp50Ala variant was associated with mild intellectual disability and speech difficulties, without motor delay or reported autistic features^22^. Its distinct functional effects contrast with those observed for variants in our cohort, many of which arose de novo, despite the X-linked inheritance of CSTF2. These findings suggest that variant-specific effects on polyadenylation may influence the severity and spectrum of neurodevelopmental manifestations.

Our RNA-seq analysis showed strongest dysregulation of polyadenylation by Phe19Leu and Gln543ter and Leu555Ser variants. While Met179Arg and Gly40Arg showed no significant and very mild effect on polyadenylation, respectively, caution should be applied in their classification as overexpression assays can naturally mask the effects of destabilising variants. In line with this, both these variants are predicted to be destabilising by the FoldX algorithm (Met179Arg ΔΔG = 2.96 and Gly40Arg ΔΔG = 3.34) requiring further experiments to understand their potential pathogenicity. Importantly, the individual with the Met179Arg variant, with autism spectrum disorder and mild intellectual disability, also carried a 16p11.2 deletion (arr[hg19]16p11.2(29656684_30190568)x1), consistent with a recurrent proximal (BP4–BP5) deletion in individuals with variably penetrant neurodevelopmental phenotypes, including intellectual disability, speech delay, learning difficulties, motor coordination abnormalities and behavioural phenotypes^36–38^. While we cannot exclude that the *CSTF2* variant may be contributing to his features, these may be attributable to the 16p11.2 microdeletion.

Overexpression of Phe19Leu and Gln543ter and Leu555Ser variants suppresses activity of hundreds of intronic polyadenylation sites and activates hundreds of polyadenylation sites downstream in 3’UTRs of corresponding genes. Based on significantly higher GU-content downstream of suppressed sites we propose that inactivation of CSTF2 directly suppresses intronic sites (and a small fraction of suppressed sites in 3’UTR) while downstream activation is likely a secondary effect. This is also supported by higher predicted strength of the suppressed sites. Previous reports demonstrated the role of CSTF2 in activation of intronic polyadenylation in immune related genes^16,39^; our results highlight CSTF2 as a global regulator of intronic polyadenylation. Importantly, we detected a strong suppressive effect on a *CSTF3* intronic site known to function as a sensor in an evolutionary conserved feedback autoregulatory loop controlling the state of polyadenylation in cells^31^.

Strikingly, the observed suppressive effect of these variants on polyadenylation was stronger and opposite from the moderate activating effect of overexpression of the Asp50Ala variant. While the degree of dysregulation by investigated variants is likely dependent on the state of polyadenylation in a cell (overall expression and activity of polyadenylation factors), the directionality of dysregulation is expected to be constant. Therefore, our data strongly suggest “gain-of-function” effect of Asp50Ala in line with previously published *in vitro* experimental data^22^. More importantly these data suggest that CSTF2 activity and/or expression tightly regulates polyadenylation in a balanced manner with both activating and inactivating variants inducing intellectual disability. Nevertheless, we would expect loss-of-function variants to occur more frequently than gain-of-function variants, as random genetic variation is more likely to disrupt existing protein function than to confer enhanced or novel activity^40,41^.

Taken together our data suggest that CSTF2 Phe19Leu and Gln543ter and Leu555Ser variants are likely inactivating “loss-of-function” variants. Together with additional ACMG/AMP criteria^42^, we suggest these variants can be classified as “Likely Pathogenic” [PS3, PM2, PP3, PP2, PM6 (in 2 of 3)]. Overall, our data strengthens the association between *CSTF2* and intellectual disability, paving the way for increasing the diagnostic yield.

Recessive loss-of-function variants in the core polyadenylation factors CLP1 and CPSF3 cause severe neurodevelopmental disorders characterized by global developmental delay, microcephaly, and brain malformations^43,44^. Our data suggest that CSTF2 deficiency results in a considerably milder phenotype. This potentially reflects the distinct specificity of these proteins: in our model CSTF2 regulates polyadenylation at a subset of sites with prominent GU-rich downstream tracts, whereas CLP1 and CPSF3 are likely more broadly required for cleavage and polyadenylation of most pre-mRNAs. Consequently, CSTF2 loss is expected to have more transcript-specific effects rather than causing a global defect in mRNA 3′-end processing (with a similar intronic to 3’UTR polyadenylation shift observed after overexpression of a pathogenic CLP1 variant in a neuronal model^45^). The milder phenotype may also be explained by partial functional compensation from its paralogue, CSTF2τ^46^. Further studies in disease-relevant models are needed to determine how defects in polyadenylation drive distinct neurodevelopmental disorders, and why disruption of different components of the polyadenylation machinery results in variable clinical severity.

Taken together, our work provides the first comprehensive characterisation of inactivating CSTF2 variants, and establishes *CSTF2* as a conclusive X-linked intellectual disability gene. Our findings significantly expand the spectrum of polyadenylation disrupting disorders underlying intellectual disability, highlight the role of CSTF2 in regulation of intronic polyadenylation and suggest an important role in human neurodevelopment.

## Methods

### Plasmid Constructs

All plasmids were propagated in the NEB 5-alpha *E. coli* strain (NEB, cat# C2987H). Human CSTF2 ORF was amplified from the pCMV-SPORT6-CSTF2 plasmid (Horizon Discovery, # MHS6278-202758031) using KAPA HiFi DNA polymerase HotStart ReadyMix (Roche, cat# KK2601) and the following primers: forward, 5’-AGAGTCGACAGAGCCATGGCGGGTTTGACTGTG-3’; and reverse, 5’-GCTTGTACAACCTATCAAGGTGCTCCAGTGGATTTCTG-3’; purified, digested with SalI and BsrGI and cloned into pEGFP-N3 (Clonetech) digested with the same restriction enzymes. All *CSTF2* variants were introduced using a modified Quikchange site-directed mutagenesis protocol, using the KAPA HiFi DNA polymerase HotStart ReadyMix (Roche, cat# KK2601) and the corresponding primers sequences provided in Supplementary Table 2. To introduce the V5 tag at N-terminus of CSTF2 ORF V5 tag sequence was ordered as two single-strand oligos: forward, 5’-GATCCATGGGCAAGCCCATCCCCAACCCCCTGCTGGGCCTGGACAGCACCGGAG-3’; and reverse, 5’-TCGACTCCGGTGCTGTCCAGGCCCAGCAGGGGGTTGGGGATGGGCTTGCCCATG-3’, (Integrated DNA Technologies, Inc.) annealed at 95°C for 5 minutes with gradual slow cooling to 25°C for and then ligated into corresponding CSTF2 pEGFP-N3 plasmids cut with SalI and XhoI.

Constructs were verified by dideoxynucleotide Sanger (Source BioScience, UK) or/and long-read sequencing (FullCirlce, UK). Plasmid and plasmid maps are available on request.

### Cell culture and transfection

HEK-293T A2 cells^47^ were kindly provided by Eugene Makeyev. Cell morphology was assessed by light microscopy and tested negative for mycoplasma contamination using the MycoGenie Rapid Detection Kit (AssayGenie, Cat# MORV0011). Cells were maintained in DMEM (Thermo Fisher Scientific, Cat# 11360070) supplemented with 10% FBS (Hyclone, Cat# SV30160.03) at 37 °C with 5% CO_2_. For passaging, cells were washed with PBS and dissociated with 0.05% Trypsin-EDTA (Thermo Fisher Scientific, Cat# 15400054) for 10 minutes at 37 °C.

For bulk RNA-sequencing and Immunoblotting cells were seeded at a density of 3×10^5^ per well of a 12-well plate. in 1 ml of culture medium containing 1 μg/ml doxycycline (Dox; Sigma, cat# D9891-1G) per well and cultured overnight. The next morning cells were transfected with 1 µg of plasmid DNA using jetOPTIMUS® DNA Transfection Reagent (Sartorius, #101000051) per manufacturer’s protocol. For 3’end proximal RNA-sequencing, cells were seeded overnight at a density of 1.5×10^5^ per well of a 24-well plate in 0.5 ml of culture medium containing 1 μg/ml doxycycline (Dox; Sigma, cat# D9891-1G) and transfected with 0.5 µg of plasmid DNA the next morning.

For RNA sequencing cells were washed with PBS and harvested 48 hours after transfection and processed as follows: for bulk RNA-seq, total RNA was purified using the RNeasy Mini Kit (Qiagen, #74104) according to the manufacturer’s protocol; for 3’end proximal RNA-sequencing, cells were collected in 100 μL Zymo DNA/RNA Shield (Zymo Research, #R1100-50) and submitted for processing to the service provider.

For western blot analysis, 48 hours after transfection cells were washed with DPBS (ThermoFisher Scientific, #14190144), and lysed in Pierce IP lysis buffer (ThermoFisher Scientific, #87788) supplemented with Halt™ Protease Inhibitor (ThermoFisher Scientific, #78430). Cell lysates were incubated on ice at 4 °C for 30 min at the bench before centrifugation at 4 °C for 15 min. The soluble protein fraction was collected, and protein concentrations were quantified using the Pierce™ BCA Protein Assay Kit (ThermoFisher Scientific, #23225).

### Immunoblotting

Immunoblotting was performed as described in ref^48^. After transfer membranes were blocked in 5% milk-TBST for 1 h, sectioned according to the molecular weights of the target proteins, and incubated for 2 h at room temperature with primary antibodies against V5 (Bio-Rad, #MCA1360, 1:1000) or ERK1/2 (loading control; Cell Signaling Technology, #9102, 1:1000) diluted in 5% milk-TBST. After three washes with TBST, membranes were incubated for 1 h at room temperature with HRP-conjugated anti-rabbit IgG (Jackson ImmunoResearch, #111-035-045-JIR-1.5 ml, 1:10000) or anti-mouse IgG (Jackson ImmunoResearch, #115-035-062-JIR-1.5 ml, 1:10000) secondary antibodies. Signal was detected using ECL Prime Western Blotting Detection Reagent (Merck, #RPN2236) on an iBright FL1000 imaging system. Image quantification was performed using Ibright analysis software (ver. 5.4.0).

### RNA-sequencing

For total RNA-sequencing library construction and sequencing were performed at Biomarker Technologies (BMKGENE) GmbH. The concentration and integrity of the extracted RNA was assessed using the Nanodrop2000 (Thermo Fisher Scientific), and LabChip GX (Perkin Elmer), respectively. The poly(A)-containing mRNA molecules were purified using poly(T) oligo-attached magnetic beads. For library preparation, the VAHTS Universal V8 RNA-seq Library Prep Kit for Illumina (NR605, Vazyme) was used, following the manufacturer’s recommendations. Library quality control was performed using the QSep-400 (BiOptic, Taipei, China) and Qubit 3.0 (Thermo Fisher Scientific). The qualified library was sequenced on the Illumina Novaseq X platform (Illumina,) with paired-end 150 bp (PE150) mode.

For 3′-end-proximal RNA sequencing, cells collected in DNA/RNA Shield were processed and sequenced by Plasmidsaurus using their 3′-end RNA-seq workflow. Total RNA was isolated using a bead-based extraction method and quantified using a fluorescence-based assay. Messenger RNA was reverse-transcribed using a poly(dT)VN primer, with unique molecular identifiers (UMIs) incorporated during first-strand cDNA synthesis to label individual RNA molecules. Following second-strand synthesis, cDNA was tagmented, amplified, and indexed with unique dual indices. Libraries were sequenced on an Illumina NovaSeq X Plus platform using asymmetric paired-end sequencing. The resulting strand-specific mRNA reads were approximately 90 bp in length and enriched towards transcript 3′ ends.

### Bioinformatics

Total RNA-sequencing reads were aligned with HISAT2 using the hg38 UCSC human genome files, converted to BAM format using SAMtools and to BEDGRAPH format using Bamtools. Differential polyadenylation site usage analyses were performed with IPAFinder and DaPars^5,30^ packages. For intronic polyadenylation site analysis with IPAFinder custom annotation (https://github.com/ZhaozzReal/IPAFinder/blob/master/anno_files/IPAFinder_anno_hg38.txt) was used to detect intronic polyadenylation sites and quantify their usage (intronic poly(A) site usage index (IPUI)) with IPAFinder_DetectIPA.py script. The outputs for each sample were used to infer statistically differential usage of the sites with Infer_DUIPA.R. For 3’UTR polyadenylation sites analysis annotation of 3’UTRs were generated using DaPars_Extract_Anno.py and UCSC annotation of 3’UTRs of hg38 human genes (hg38.knownGene.gtf). Generated annotation and aggregated bedgraphs for analysed samples were used to infer percentage of distal poly(A) site usage index (PDUI) and statistically significant changes with DaPars_main.py.

3′-end-proximal RNA-seq data (BAM files, quality filtered, trimmed, aligned to GRCh38 with STAR aligner and deduplicated using UMIs by Plasmidsaurus) were mapped to the region 500bp upstream of all annotated polyadenylation sites from Polya_DB version 4.1^49^ (https://exon.apps.wistar.org/polya_db/v4/). For sites located closer than 250bp with similar pattern of usage (Pearson correlation coefficient for number of reads across all samples >0.95) only the site with higher number of mapped reads was kept for further analysis. Sites with low number of reads were filtered out using *filterByExpr* function form edgeR package^50^ followed by trimmed mean of M-values (TMM) normalisation and voom transformation. Linear models were fitted for each site, and pairwise comparisons between individual *CSTF2* variants and wild-type *CSTF2* were performed using limma^51^. Differential polyadenylation-site usage was assessed using *diffSplice*, comparing the log₂ fold change of each site with the weighted average change of other sites within the same gene. Statistical significance was determined using empirical Bayes moderated t-tests, with P values adjusted using the Benjamini–Hochberg method separately for each comparison.

Percentage of GU nucleotide was calculated in 50bp window downstream of annotated polyadenylation site. Evolutionary conservation was calculated as a mean of phastCons100way.UCSC.hg38 score in 100bp window centred at all annotated polyadenylation sites using GenomicScores R package^52^. Strength of polyadenylation site measurements were taken from Polya_DB version 4.1 (“PolyaStrength” column).

Genomic intervals were analysed using Bedtools or custom R-scripts. Duplicated features with identical genome positions and gene names were removed from the analyses.

### Structural modelling

Models of the RRM domain of human CSTF2 in complex with a UGUGUUG RNA oligomer were created using the Alphafold3 server and default parameters. The reported, scaled, pLLDT values were obtained directly from the output models. The models were then visualized using Pymol^53^

### Statistical analysis

Statistical analyses were performed in R (v4.4.1). All samples analysed at least in biological duplicates. Group comparisons were performed using two-sided Wilcoxon rank-sum, One way ANOVA with Tukey’s post hoc, Fisher’s exact tests, as appropriate. Multiple testing was controlled using the Benjamini–Hochberg method, and adjusted *P* < 0.05 was considered statistically significant.

### Patients and ethics

This work used data from the 100 000 Genomes Project (IRAS ID 166046; East of England - Cambridge South Research Ethics Committee reference 14/EE/1112), which obtained written informed consent from all participants (or from their parent/legal guardian). All investigations were conducted in accordance with the tenets of the Declaration of Helsinki.

This work used data from the National Genomic Research Library [East of England - Cambridge Central Research Ethics Committee reference 20/EE/0035].

This work was conducted under the registered and approved Genomics England Clinical Research project ID RR1011.

The SPARK study is funded by the Simons Foundation and uses a single, central Institutional Review Board (WCG IRB Protocol #20151664). Informed consent was obtained by the SPARK study team from the participant or the caregiver/parent.

Written informed consent from patient or legal guardian were obtained for publication of individual data.

The criteria application and variant classification presented in this paper does not necessarily reflect the criteria and classification of the clinical testing laboratories involved in this paper.

## Data availability

Data from the National Genomic Research Library (NGRL) used in this research are available within the secure Genomics England Research Environment. Access to NGRL data is restricted to adhere to consent requirements and protect participant privacy. Data used in this research include: variant tiering and clinical phenotype data from 100kGP Clinical (release v19 31.10.2024) and NHS Genomic Medicine Service (GMS, release v5 25.08.2025). No additional datasets were generated.

Access to NGRL data is provided to approved researchers who are members of the Genomics England Research Network, subject to institutional access agreements and research project approval under participant-led governance. For more information on data access, visit: https://www.genomicsengland.co.uk/research

Approved researchers can obtain the SPARK population dataset described in this study (SPARK Consortium iWES v3 dataset) by applying at https://base.sfari.org. SPARK data were accessed via SFARI Base and Genotypes and Phenotypes in Families (GPF) platform^54^. The de-identified SFARI subject IDs have been retained in accordance with the SFARI Researcher Distribution Agreement, under which data must be indexed using official subject IDs to allow findings to be linked across studies without names or other direct personal identifiers being disclosed.

The sequencing data generated during this study will be deposited in the European Genome-phenome Archive prior to journal peer-reviewed publication. Until then, data are available from the corresponding author upon reasonable request, subject to data-sharing agreements.

## Acknowledgements

This study has been possible thanks to a grant from Guy’s C St Thomas’ Charity grant STR111001. This research was in part supported by Royal Society grant RG\R1\251638 and Friends of Guy’s and St Thomas’ Hospital Research Grant 24/02R. AR is funded by UCL and MRC grant Z504695/.1

We gratefully acknowledge the participants of the National Genomic Research Library (NGRL), whose contributions made this research possible. Secure access to the NGRL under project ID RR1011 was provided by Genomics England, which delivers the NGRL in partnership with NHS England, and is wholly owned by the UK Department of Health and Social Care. The NGRL contains participants’ health data collected by the NHS as part of their care, along with samples and data from their participation in research, for which fully informed consent has been obtained. This includes genomic and clinical data provided through the NHS Genomic Medicine Service, as well as data obtained through research studies, including the 100,000 Genomes Project and the Generation Study, both of which are delivered in partnership with the NHS, and from other research cohorts involving external collaborators.

We are grateful to all of the families in SPARK, the SPARK clinical sites and SPARK staff. We appreciate obtaining access to phenotypic and genetic data from SPARK Consortium iWES v3 dataset on SFARI Base and GPF viewer by Simons Foundation.

We thank Eugene Makeyev, Suzi Walker, Genomics England clinical collaboration team and Anna Zhuravskaya for advice, insightful discussions, help and support.

## Disclosure/Conflict of interests

Ingrid M. Wentzensen is an employee of and may own stock in GeneDx.

---

**Supplementary figure:**
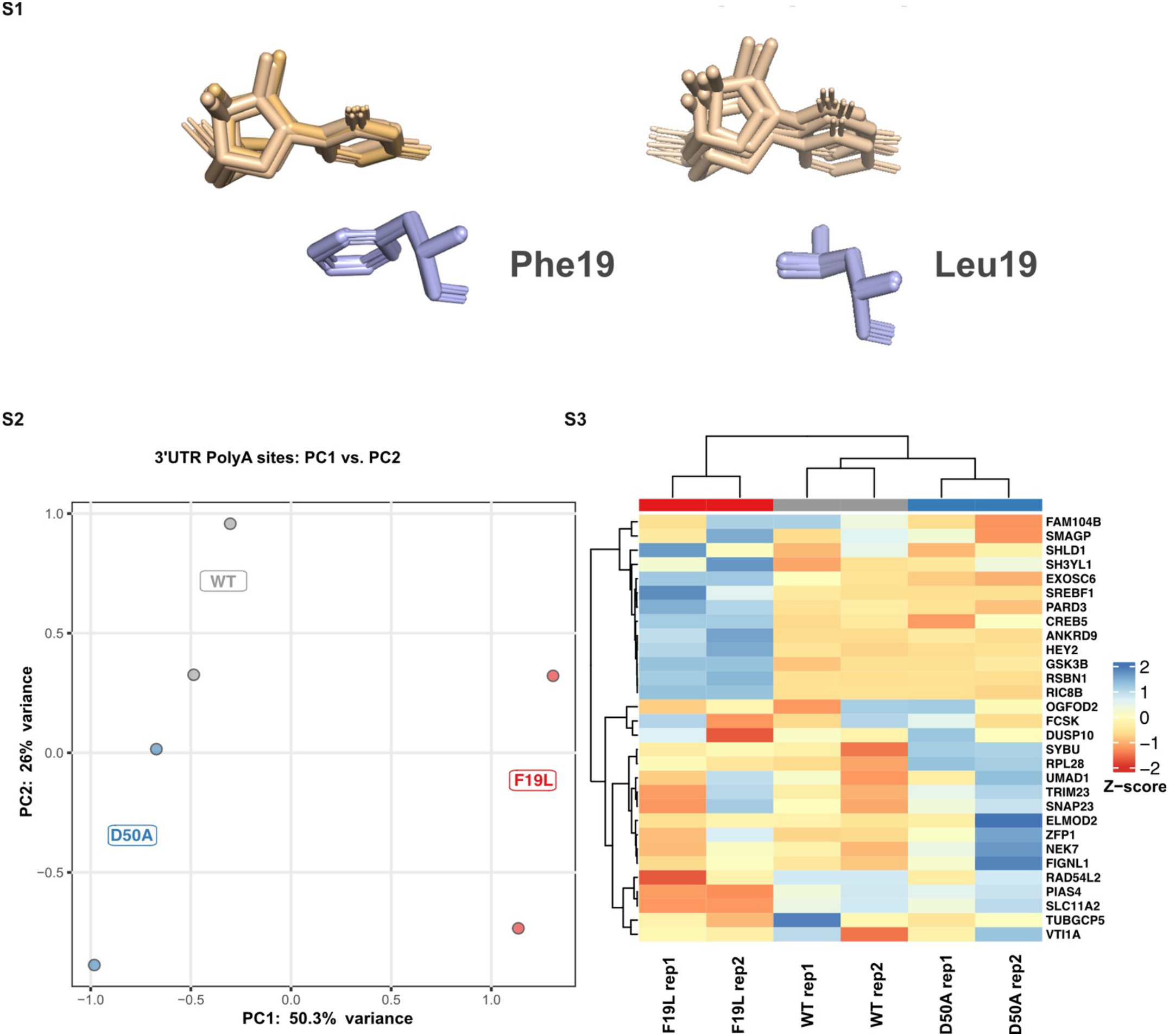
**S1** – (*Left*) In all five Alphafold3 models of wild-type protein in complex with the cognate RNA, the aromatic side chain in position 19 is stacked on the closest RNA base. (*Right*) In all the equivalent models of the Phe19Leu mutant, this contact is lost. Please note the Pymol superimposition of the models displayed here is across the whole structure. Notably, in the wild-type protein, the normalized per-atom confidence estimate (pLDDT) for Phe19 varies between 92 and 96, while that of the stacked nucleotide varies between 80 and 86. In the equivalent groups in the mutant’s model, the corresponding scores vary between 86-96 for the protein, and 78 and 84 for the nucleotide. A slightly lower score for the nucleotide is not unexpected, given the relatively dynamic nature of the protein-RNA interactions and that the confidence of the ligand is calculated with respect to the protein chain. **S2** - PCA of 100 most variable 3’UTR polyadenylation events significantly dysregulated by overexpression of either Asp50Ala CSTF2 or Phe19Leu CSTF2 in comparison with WT CTSF2 in HEK 293T A2 cells (DAPARS centred and unscaled PDUI values, BH adjusted p < 0.05). Samples were projected onto the first two principal components, with group centroids calculated as the mean PC coordinates per condition. **S3** - Heatmap of the top 30 most variable 3’UTR polyadenylation events significantly dysregulated by overexpression of either Asp50Ala CSTF2 or Phe19Leu CSTF2 in comparison with WT CTSF2. Rows and columns were hierarchically clustered using correlation distance and Ward’s D2 linkage (pheatmap, R). PDUI values were scaled by row Z-score and visualized with a red–white–blue colour scheme.

**Supplementary Table 2:**
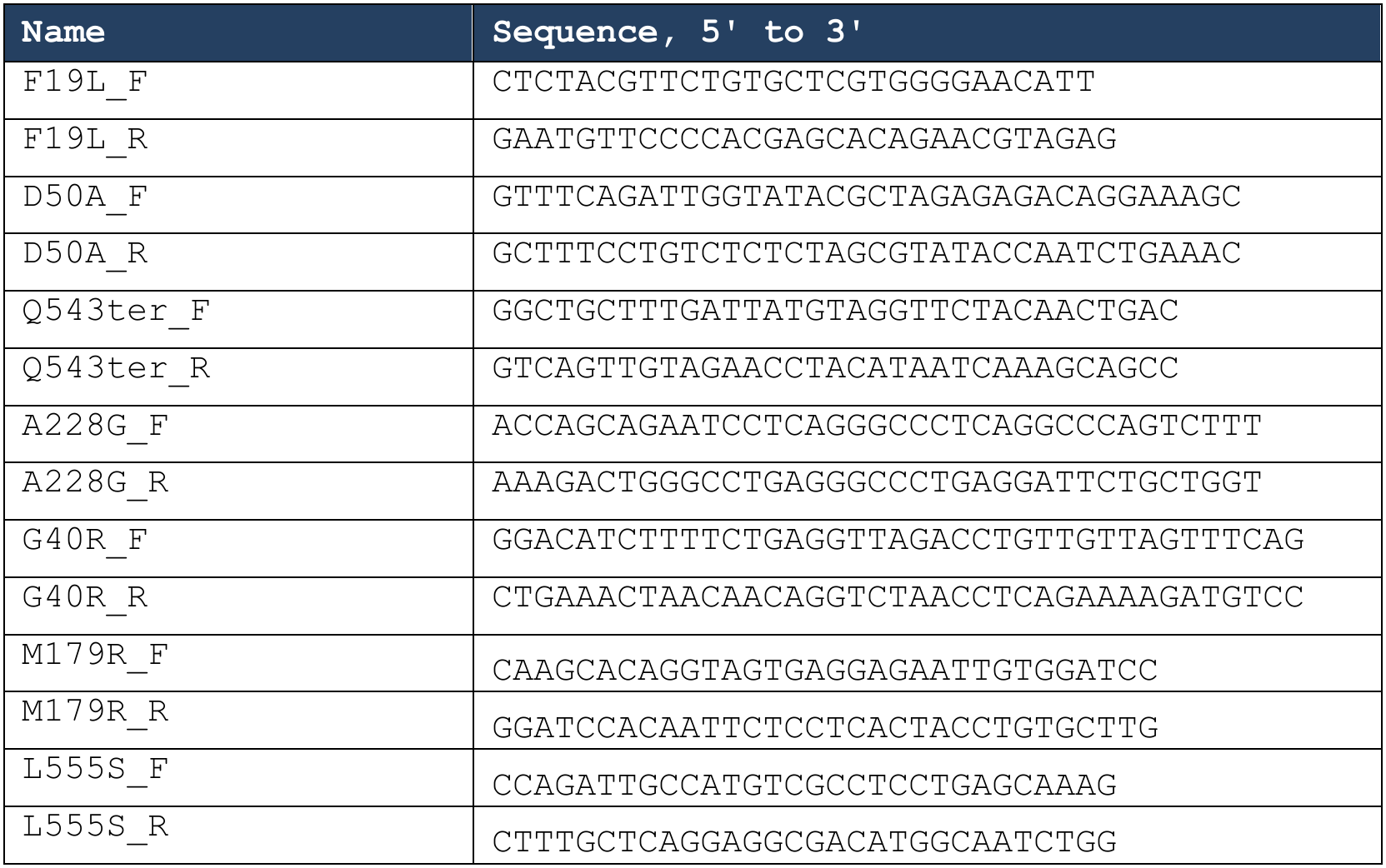
Oligonucleotides used for site-directed mutagenesis.

## Notes

### Author Declarations

This work used data from the 100 000 Genomes Project (IRAS ID 166046; East of England - Cambridge South Research Ethics Committee reference 14/EE/1112), which obtained written informed consent from all participants (or from their parent/legal guardian). All investigations were conducted in accordance with the tenets of the Declaration of Helsinki. This work used data from the National Genomic Research Library [East of England - Cambridge Central Research Ethics Committee reference 20/EE/0035]. This work was conducted under the registered and approved Genomics England Clinical Research project ID RR1011. The SPARK study is funded by the Simons Foundation and uses a single, central Institutional Review Board (WCG IRB Protocol #20151664). Informed consent was obtained by the SPARK study team from the participant or the caregiver/parent. Written informed consent from patient or legal guardian were obtained for publication of individual data. The criteria application and variant classification presented in this paper does not necessarily reflect the criteria and classification of the clinical testing laboratories involved in this paper.

